# Medical Costs, Health Care Utilization, and Productivity Losses Associated With Hypertension by COVID-19 Among US Commercial Enrollees

**DOI:** 10.1101/2024.05.31.24308307

**Authors:** Jun Soo Lee, Yidan (Xue) Zhang, Yu Wang, Joohyun Park, Ashutosh Kumar, Bruce Donald, Feijun Luo, Kakoli Roy

**Author notes:** Feijun Luo and Kakoli Roy are co-senior authors. Corresponding author: Jun Soo Lee, PhD, Division for Heart Disease and Stroke Prevention Centers for Disease Control and Prevention, 4770 Buford Hwy, Building 107, Atlanta, GA 30341. Disclaimer: The findings and conclusions in this article are those of the authors and do not necessarily represent the official position of the Centers for Disease Control and Prevention. Use of trade names and commercial sources is for identification only and does not imply endorsement by the US Department of Health and Human Services. There is no potential conflict of interest related to any part of this article. Sources of funding: The authors received no financial support for the research, authorship, and/or publication of this article.

## Abstract

**Background:** Hypertension is a major risk factor for cardiovascular and renal diseases, significantly contributing to morbidity and mortality. The COVID-19 pandemic has heightened concerns about the impact of hypertension on severe COVID-19 outcomes.

**Methods:** We conducted a cross-sectional analysis using the 2021 MarketScan Commercial and Health and Productivity Management databases. The study included adults aged 18-64 with continuous employer-sponsored private insurance, excluding those with pregnancy or capitated plans. We compared excess total medical costs, healthcare utilization (including the number of emergency department visits, inpatient admissions, outpatient visits, and outpatient prescription drugs), and productivity losses and related costs due to sick absences, short-term disability (STD), and long-term disability (LTD) between individuals with and without hypertension, further stratified by COVID-19 diagnosis. Multivariate regression models adjusted for demographics and comorbidities were used to estimate the differences in outcomes.

**Results:** Among 1,612,398 adults aged 18-64 years, 13% had hypertension in 2021. Those with hypertension were older, were less likely to be female or live in urban areas, and exhibited a higher prevalence of comorbidities. The total excess medical costs associated with hypertension were $8723 per patient (95% CI, $8352-$9093), which was significantly higher by $6117 (95% CI, $4780-$7453) among individuals diagnosed with COVID-19. Persons with hypertension had higher health care utilization, including a higher number of ED visits (0.21 per patient; 95% CI, 0.21-0.22), inpatient admissions (0.11; 95% CI, 0.10-0.12), outpatient visits (5.42; 95% CI, 5.36-5.49), and outpatient prescription drugs (10.85; 95% CI, 10.75-10.94). Moreover, they experienced a greater number of sick absences (1.22 days; 95% CI, 1.07-1.36) and STD occurrences (3.68 days; 95% CI, 3.38-3.98) per patient compared to those without hypertension. These trends were further exacerbated among individuals diagnosed with COVID-19.

**Conclusions:** Hypertension markedly increases medical costs, healthcare utilization, and productivity losses, which are further exacerbated by COVID-19. These findings highlight the substantial economic burden of managing hypertension in the context of the COVID-19 pandemic and underscore the importance of targeted interventions.

## INTRODUCTION

Hypertension (ie, high blood pressure [BP]) is a major risk factor of cardiovascular and renal diseases, which are among the leading contributors of all-cause mortality.^1,2^ Recent national estimates indicate that approximately 122 million (47%) adults aged 20 years and older have hypertension in the United States (US).^3,4^ Moreover, hypertension has been consistently linked to elevated health care utilization and increased medical cost.^5–8^ For instance, in 2019, hypertension was implicated in over 18 million hospital discharges, with 1.4 million cases wherein hypertension was the primary diagnosis.^3,9^ The estimated medical cost associated with hypertension was reported to be $1500 per person per year in 2012-2013, contributing to a staggering $109 billion (in 2015 US dollars) in total annual medical cost nationwide.^6^

In addition to direct medical costs, hypertension is associated with productivity loss among working-age adults.^10^ Hypertension often goes unnoticed and remains untreated or uncontrolled, indicating BP levels persistently above the recommended thresholds,^11^ among working-age adults.^12^ For example, 93.4% and 74.4% of hypertensive adults in the US aged 18-44 and 45-64 years had uncontrolled hypertension, respectively, compared with 69.7% of adults aged 65 years and above.^13^ Uncontrolled hypertension is associated with comorbidities that can lead to productivity loss. A systematic literature review of studies published from 1990 through 2015 suggests that hypertension-associated absenteeism, presenteeism, and short-term disability cost upwards of $60, $70, and $50 per person per year, respectively, resulting in billions of dollars in total annual productivity loss nationwide (eg, $11 billion from absenteeism alone).^10,14–16^

In parallel, emerging evidence following the onset of the COVID-19 pandemic has pointed to greater morbidity and mortality among COVID-19 patients with comorbid hypertension.^17–19^ However, research also indicates that hypertension may independently affect COVID-19 prognosis through heightened immunological activity and organ damage, primarily in those with uncontrolled systolic BP.^20,21^ Compared with peers without hypertension, COVID-19 patients with hypertension are significantly more likely to be hospitalized and require treatment in the intensive care unit.^17,18^ Also, COVID-19 infections may lead to the onset of hypertension in people without preexisting hypertension.^22^

Despite this growing body of evidence, a significant gap remains in understanding the interaction between hypertension and COVID-19,^23–25^ especially regarding the impact on medical costs, health care utilization, and productivity losses.^26–29^ To bridge this gap, our study aims to investigate the economic burden of hypertension during the COVID-19 pandemic, specifically focusing on medical costs, health care utilization, and productivity losses among employed adults aged 18-64 years with employer-sponsored private insurance. Additionally, the study seeks to delineate variations in hypertension-related medical costs and productivity losses based on COVID-19 diagnosis status. By addressing these gaps, our study may elucidate the intertwined effects of COVID-19 and hypertension on total medical costs, health care utilization, and productivity loss.

## METHODS

### Data

We used the MarketScan Commercial and Health and Productivity Management (HPM) databases from 2021.^30^ The MarketScan Commercial Database contained person-level paid claims and encounters, including medical costs (patients’ out-of-pocket costs and insurance payments) and information on health care utilization, such as inpatient services, outpatient services, emergency department (ED) visits, and outpatient prescription drug claims. The data came from approximately 350 payers, encompassing over 20 billion service records provided by

>300 employers, >30 health plans, and >500 hospitals nationwide.^30^ The HPM Database contained detailed payroll-related information about employees, including sick absences, short-term disability (STD), and long-term disability (LTD). All data were de-identified, and the study was exempt from review by the Institutional Review Board of the Centers for Disease Control and Prevention.

### Identification of Patients With Hypertension

Our sample included employed individuals aged 18-64 years who were continuously enrolled in an employer-sponsored insurance plan throughout 2021. The sample excluded individuals with a pregnancy diagnosis (codes listed in eTable 1) or with capitated insurance in 2021. Finally, we limited the analytic sample to individuals in the HPM Database, who had complete information on absences, STD, or LTD in 2021. Because not all employers provided payroll information, we stratified the sample by individuals with medical cost, sick absence, and STD or LTD information. In each stratified subsample, we identified individuals with hypertension if they had at least 1 inpatient or ED visit or 2 outpatient visits containing a hypertension diagnosis (International Classification of Diseases, 10th Revision, Clinical Modification [ICD-10-CM] I10 through I15) in 2021 (eTable 1).

### Outcome Variables

The primary outcome variables, measured in per person per year, are (1) total medical costs (sum of patients’ out-of-pocket costs and insurance payments), (2) health care utilization, including the number of ED visits, inpatient admissions, outpatient visits, and outpatient prescription drugs, (3) total missed workdays related to sick absences, STDs, and LTDs, and (4) productivity costs calculated on total missed 8-hour workdays due to sick absences, STDs, and LTDs in 2021.

Estimated employer productivity costs were computed by multiplying the number of hours absent from the number of missed workdays times 8 hours, with the average hourly wage in 2021.^31^ We adjusted for absences associated with STD (70% of hourly wage) and LTD (60% of hourly wage).^31,32^ The average hourly wage ($30.61) for all employees in private nonfarm payrolls was obtained from the 2021 US Bureau of Labor Statistics (BLS).^33^

### Independent Variable

The primary independent variable is a binary variable indicating whether individuals had hypertension in 2021. We interacted this variable with a binary indicator for COVID-19 diagnosis (individuals with ICD-10-CM of U07.1 in 2021) to examine the interaction between hypertension and COVID-19 with respect to the outcome variables.

### Control Variables

All models were adjusted for age groups (aged 18-34 [reference], 35-44, 45-54, and 55-64 years), sex (female [reference], male), residency (rural [reference] and urban),^34^ census region (Northeast [reference], Midwest, South, and West), and comorbidities. Comorbidities were defined using 17 conditions outlined in the Quan Charlson Comorbidities.^35^ Each comorbidity was identified if individuals had ≥1 inpatient admission or ≥2 outpatient encounters (≥30 days apart) using the ICD-10-CM diagnosis codes as specified in Quan et al.^35^

### Statistical Analysis

We first documented summary statistics of patients’ characteristics, including age, sex, rural-urban residence, census region, and comorbidities. We tested the differences in average values by hypertension diagnosis status using the Wilcoxon rank-sum test for continuous variables and Pearson’s chi-square test for categorical variables.

In the regression analyses, where the dependent variables were total medical costs and productivity costs (related to sick absences, STD, and LTD), we used a generalized linear model (GLM) with a log-link function and gamma distribution. For dependent variables that were count variables, specifically outcomes related to health care utilization (including the number of ED visits, inpatient admissions, and outpatient prescription drugs) and missed workdays (due to sick absences, STD, and LTD), a negative binomial regression model was used. We ran separate regressions for each dependent variable to estimate the association with hypertension status (with and without hypertension) as well as the association including the interaction of hypertension status and COVID-19 diagnosis (with and without COVID-19). The remaining control variables were the same for all models. We reported the predicted outcomes variables in terms of average marginal effects (AMEs) for individuals, both with and without hypertension, and for those with and without COVID-19. Furthermore, we documented the differences in the estimated AMEs by combined hypertension status and by COVID-19 diagnosis status. AMEs represent the change in the predicted outcome variables associated with a one-unit change in the independent variable, holding all other variables constant. AMEs provide estimates of the predicted changes in outcomes resulting from variations in hypertension and COVID-19 diagnosis. We defined the differences in AMEs associated with hypertension and COVID-19 diagnosis status as “excess” outcomes associated with hypertension and COVID-19.

In the sensitivity analysis, we applied a zero-inflated negative binomial regression model for the count variables—specifically missed workdays attributable to sick absences, STD, and LTD—and a two-part model for productivity costs due to potential excess zero values in the dependent variables. In the two-part model, we first used a logistic regression to estimate the probability of non-zero expenditures, followed by GLM regression with log-link in the second part.

All analyses used 2021 data, which was the most recently available data. Also, there might have been interruptions to medical care during the early COVID-19 pandemic in 2020,^36,37^ as the severity of COVID-19 and its relationship with hypertension might have been different during the early and middle stages of the pandemic. To test this potential interruption, we conducted additional sensitivity analyses using 2020 data. The sensitivity analysis results using 2020 were adjusted to 2021 dollars using the Consumer Price Index from the US BLS.^38^

Two-tailed tests with *P* values of <0.05 were used to indicate statistical significance. All analyses were conducted using Stata SE version 17 (StataCorp, College Station, Texas).

## RESULTS

We identified 1 612 398 employed individuals aged 18-64 years who were continuously enrolled in a noncapitated employer-sponsored private insurance plan without a pregnancy history in 2021 (Figure 1). Among these individuals, 139 490 (9%) had non-missing information on workplace absences, 1 355 064 (84%) had non-missing information on STD, and 1 331 963 (83%) had non-missing information on LTD. A total of 215 480 (13%) individuals had hypertension in 2021. Persons with hypertension were older (51.2 vs 42.9 years; *P* < 0.001), were less likely to be female (34.0% vs 39.4%; *P* < 0.001), were less likely to reside in urban areas (90.4% vs 92.7%; *P* < 0.001), and had a higher likelihood of having comorbidities (*P* < 0.001 in all comorbidities) (Table 1).

**Figure 1.**
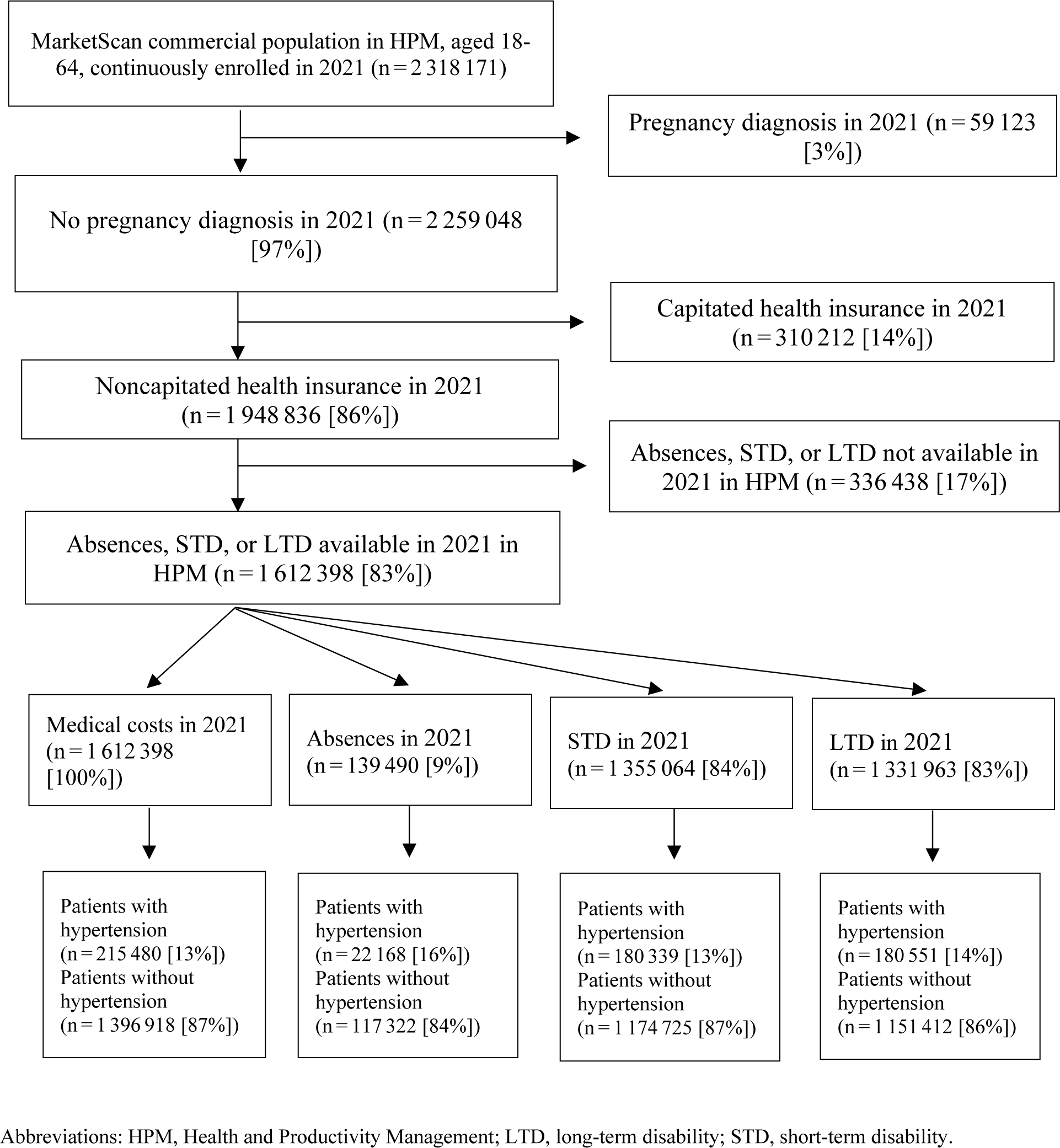
Study Sample Selection of Patients Diagnosed With Hypertension, MarketScan® Commercial and Health and Productivity Management Database, 2021

**Table 1.** Summary Statistics, MarketScan® Commercial Insurance, 2021^a^.

|  | All | Without hypertension | With hypertension |
| --- | --- | --- | --- |
|  | N = 1 612 398 | N = 1 396 918 | N = 215 480 |
| Age, mean (SD) | 44.0 (11.1) | 42.9 (11.0) | 51.2 (8.5)*** |
| **Age groups, n (%)** |  |  |  |
| 18-34 | 394 617 (24.47%) | 383 744 (27.47%) | 10 873 (5.05%)*** |
| 35-44 | 411 201 (25.50%) | 376 307 (26.94%) | 34 894 (16.19%)*** |
| 45-54 | 451 159 (27.98%) | 372 585 (26.67%) | 78 574 (36.46%)*** |
| 55-64 | 355 421 (22.04%) | 264 282 (18.92%) | 91 139 (42.30%)*** |
| Female, n (%) | 623 335 (38.66%) | 550 081 (39.38%) | 73 254 (34.00%)*** |
| Urban residency, n (%) | 1 489 709 (92.39%) | 1 294 879 (92.70%) | 194 830 (90.42%)*** |
| **Census regions, n (%)** |  |  |  |
| Northeast | 239 802 (14.87%) | 211 483 (15.14%) | 28 319 (13.14%)*** |
| Midwest | 387 301 (24.02%) | 334 612 (23.95%) | 52 689 (24.45%)*** |
| South | 694 461 (43.07%) | 585 403 (41.91%) | 109 058 (50.61%)*** |
| West | 283 790 (17.60%) | 259 426 (18.57%) | 24 364 (11.31%)*** |
| **COVID-19 diagnosis, n (%)** | 144 896 (8.99%) | 118 940 (8.51%) | 25 956 (12.05%)*** |
| **Comorbidities, n (%)** |  |  |  |
| Myocardial infarction | 8 243 (0.51%) | 2 901 (0.21%) | 5342 (2.48%)*** |
| Congestive heart failure | 8648 (0.54%) | 2378 (0.17%) | 6270 (2.91%)*** |
| Peripheral vascular disease | 7538 (0.47%) | 2959 (0.21%) | 4579 (2.13%)*** |
| Cerebrovascular disease | 2538 (0.16%) | 944 (0.07%) | 1594 (0.74%)*** |
| Dementia | 119 (0.01%) | 56 (0.00%) | 63 (0.03%)*** |
| Chronic pulmonary disease | 36 105 (2.24%) | 24 416 (1.75%) | 11 689 (5.42%)*** |
| Rheumatic disease | 9912 (0.61%) | 7154 (0.51%) | 2758 (1.28%)*** |
| Peptic ulcer disease | 1322 (0.08%) | 767 (0.05%) | 555 (0.26%)*** |
| Mild liver disease | 14 064 (0.87%) | 8241 (0.59%) | 5823 (2.70%)*** |
| Diabetes without chronic complication | 91 093 (5.65%) | 41 273 (2.95%) | 49 820 (23.12%)*** |
| Diabetes with chronic complication | 18 362 (1.14%) | 6706 (0.48%) | 11 656 (5.41%)*** |
| Hemiplegia or paraplegia | 1284 (0.08%) | 554 (0.04%) | 730 (0.34%)*** |
| Renal disease | 10 807 (0.67%) | 2459 (0.18%) | 8348 (3.87%)*** |
| Any malignancy | 25 244 (1.57%) | 17 617 (1.26%) | 7627 (3.54%)*** |
| Moderate or severe liver disease | 767 (0.05%) | 372 (0.03%) | 395 (0.18%)*** |
| Metastatic solid tumor | 3396 (0.21%) | 2212 (0.16%) | 1184 (0.55%)*** |
| AIDS/HIV | 3447 (0.21%) | 2527 (0.18%) | 920 (0.43%)*** |

Abbreviations: SD, standard deviation.
<sup>a</sup> We used the Wilcoxon nonparametric rank-sum test to test the differences in means for continuous variables, and the Pearson's chi-square test to test differences in proportions for categorical variables by medication nonadherence to antihypertensive drug status. \*\*\* $P < 0.001$

Table 2 presents the total adjusted medical costs and health care utilization associated with hypertension, including by COVID-19 diagnosis. The mean total excess medical costs associated with hypertension compared to those without hypertension were $8723 per patient per year (95% CI, $8352-$9093; *P* < 0.001). The total excess medical costs associated with hypertension were significantly higher among persons with COVID-19 than those without COVID-19 ($14 290 vs $8173; the difference was $6117 [95% CI, $4780-$7453; *P* < 0.001]).

**Table 2.** Total Medical Costs and Health Care Utilization Associated With Hypertension, 2021^a^.

|  | All | Without COVID-19 diagnosis | With COVID-19 diagnosis | With COVID-19 vs without COVID-19 |
| --- | --- | --- | --- | --- |
| **Total medical costs, \$** | | | | |
| Without hypertension | 9056 | 8228 | 17 446 | 9219*** |
|  | (8724 to 9387) | (7945 to 8510) | (16 518 to 18 374) | (8519 to 9918) |
| With hypertension | 17 779 | 16 401 | 31 736 | 15 335*** |
|  | (17 131 to 18 426) | (15 830 to 16 971) | (29 801 to 33 671) | (13 718 to 16 952) |
| Difference<sup>b</sup> | **8723\*\*\*** | **8173\*\*\*** | **14 290\*\*\*** | **6117\*\*\*** |
|  | **(8352 to 9093)** | **(7831 to 8515)** | **(12 888 to 15 691)** | **(4780 to 7453)** |
| Observations | 1 612 398 | 1 467 502 | 144 896 | 1 612 398 |
| **Health care utilization** |  |  |  |  |
| **Number of emergency department visits** |  |  |  |  |
| Without hypertension | 0.176 | 0.148 | 0.461 | 0.313*** |
|  | (0.175 to 0.177) | (0.147 to 0.149) | (0.454 to 0.468) | (0.306 to 0.320) |
| With hypertension | 0.388 | 0.348 | 0.789 | 0.441*** |
|  | (0.383 to 0.392) | (0.344 to 0.352) | (0.767 to 0.811) | (0.419 to 0.463) |
| Difference<sup>b</sup> | **0.211\*\*\*** | **0.200\*\*\*** | **0.328\*\*\*** | **0.128\*\*\*** |
|  | **(0.207 to 0.216)** | **(0.195 to 0.204)** | **(0.305 to 0.351)** | **(0.105 to 0.151)** |
| Observations | 1 612 398 | 1 467 502 | 144 896 | 1 612 398 |
| **Number of inpatient admissions** |  |  |  |  |
| Without hypertension | 0.0476 | 0.0297 | 0.229 | 0.199*** |
|  | (0.0443 to 0.0508) | (0.0280 to 0.0313) | (0.208 to 0.249) | (0.180 to 0.218) |
| With hypertension | 0.156 | 0.111 | 0.604 | 0.492*** |
|  | (0.146 to 0.165) | (0.105 to 0.117) | (0.549 to 0.658) | (0.442 to 0.542) |
| Difference<sup>b</sup> | **0.108\*\*\*** | **0.0817\*\*\*** | **0.375\*\*\*** | **0.293\*\*\*** |
|  | **(0.101 to 0.115)** | **(0.0772 to 0.0861)** | **(0.337 to 0.413)** | **(0.259 to 0.328)** |
| Observations | 1 612 398 | 1 467 502 | 144 896 | 1 612 398 |
| **Number of outpatient visits** |  |  |  |  |
| Without hypertension | 8.522 | 8.223 | 11.55 | 3.329*** |
|  | (8.505 to 8.539) | (8.207 to 8.240) | (11.48 to 11.62) | (3.260 to 3.399) |
| With hypertension | 13.94 | 13.66 | 16.81 | 3.151*** |
|  | (13.88 to 14.01) | (13.60 to 13.73) | (16.61 to 17.02) | (2.937 to 3.365) |
| Difference<sup>b</sup> | **5.422\*\*\*** | **5.438\*\*\*** | **5.260\*\*\*** | -0.178 |
|  | **(5.357 to 5.486)** | **(5.370 to 5.505)** | **(5.043 to 5.477)** | **(-0.402 to 0.0467)** |
| Observations | 1 612 398 | 1 467 502 | 144 896 | 1 612 398 |
| **Number of outpatient prescription drugs** |  |  |  |  |
| Without hypertension | 9.824 | 9.596 | 12.13 | 2.536*** |
|  | (9.802 to 9.845) | (9.574 to 9.617) | (12.05 to 12.21) | (2.459 to 2.613) |
| With hypertension | 20.67 | 20.40 | 23.39 | 2.988*** |
|  | (20.58 to 20.76) | (20.31 to 20.50) | (23.09 to 23.69) | (2.677 to 3.299) |
| Difference<sup>b</sup> | **10.85\*\*\*** | **10.81\*\*\*** | **11.26\*\*\*** | **0.452\*\*** |
|  | **(10.75 to 10.94)** | **(10.71 to 10.91)** | **(10.95 to 11.57)** | **(0.132 to 0.772)** |
| Observations | 1 612 398 | 1 467 502 | 144 896 | 1 612 398 |

\*\*\* $P < 0.001$ , \*\* $P < 0.01$ , \* $P < 0.05$ .
<sup>a</sup> We used a generalized linear model for total medical costs and a negative binomial model for the number of emergency department visits, inpatient admissions, outpatient visits, and outpatient prescription drugs. All estimates were average marginal effects with 95% confidence interval (CI), adjusting patients' age groups, sex, urbanicity of residence, Census regions, and comorbidities.
<sup>b</sup> The differences in the predicted values, ie, average marginal effects associated with hypertension.

Individuals with hypertension had a higher number of ED visits (0.211 per patient; 95% CI, 0.207-0.216; *P* < 0.001), inpatient admissions (0.108; 95% CI, 0.101-0.115; *P* < 0.001), outpatient visits (5.422; 95% CI, 5.357-5.486; *P* < 0.001), and outpatient prescription drugs (10.85; 95% CI, 10.75-10.94; *P* < 0.001) than those without hypertension (Table 2). Compared with adults without COVID-19 diagnosis, those with COVID-19 had a significantly higher number of excess ED visits (0.128; 95% CI, 0.105-0.151; *P* < 0.001), inpatient admission (0.293; 95% CI, 0.259-0.328; *P* < 0.001), and outpatient prescription drugs (0.452; 95% CI, 0.132-0.772; *P* < 0.001) associated with hypertension compared to those without hypertension (Table 2).

Table 3 documents the productivity losses and costs associated with hypertension by COVID-19 diagnosis. Individuals with hypertension had a higher number of sick absences (1.215 days per patient; 95% CI, 1.068-1.362; *P* < 0.001), STD (3.680; 95% CI, 3.381-3.980; *P* < 0.001), and higher sick absence costs ($297.3 per patient; 95% CI, $254.1-$340.6; *P* < 0.001) and STD costs ($631.0; 95% CI, $591.4-$670.7; *P* < 0.001). Compared with persons without COVID-19 diagnosis, those with COVID-19 had a significantly higher number of excess STD (2.412; 95% CI, 0.837-3.987; *P* < 0.01) and STD costs ($413.6; 95% CI, $205.0-$622.2; *P* < 0.001) associated with hypertension.

**Table 3.** Productivity Losses and Costs Associated With Hypertension, 2021^a^.

|  | All | Without COVID-19 diagnosis | With COVID-19 diagnosis | With COVID-19 vs without COVID-19 |
| --- | --- | --- | --- | --- |
| **Sick Absences (days)** |  |  |  |  |
| Without hypertension | 2.181 | 1.990 | 3.870 | 1.880*** |
|  | (2.137 to 2.225) | (1.950 to 2.031) | (3.658 to 4.081) | (1.668 to 2.091) |
| With hypertension | 3.396 | 3.220 | 4.952 | 1.732*** |
|  | (3.255 to 3.536) | (3.078 to 3.361) | (4.426 to 5.478) | (1.193 to 2.270) |
| Difference<sup>b</sup> | **1.215\*\*\*** | **1.230\*\*\*** | **1.082\*\*\*** | -0.148 |
|  | **(1.068 to 1.362)** | **(1.083 to 1.377)** | **(0.514 to 1.650)** | **(-0.727 to 0.431)** |
| Observations | 139 490 | 125 345 | 14 145 | 139 490 |
| **STD (days)** |  |  |  |  |
| Without hypertension | 2.736 | 2.349 | 6.699 | 4.351*** |
|  | (2.628 to 2.844) | (2.262 to 2.435) | (6.212 to 7.186) | (3.903 to 4.799) |
| With hypertension | 6.416 | 5.815 | 12.58 | 6.763*** |
|  | (6.082 to 6.750) | (5.506 to 6.123) | (10.99 to 14.16) | (5.195 to 8.331) |
| Difference<sup>b</sup> | **3.680\*\*\*** | **3.466\*\*\*** | **5.878\*\*\*** | **2.412\*\*** |
|  | **(3.381 to 3.980)** | **(3.184 to 3.748)** | **(4.312 to 7.444)** | **(0.837 to 3.987)** |
| Observations | 1 355 064 | 1 234 579 | 120 485 | 1 355 064 |
| **LTD (days)** |  |  |  |  |
| Without hypertension | 0.842 | 0.703 | 2.236 | 1.533 |
|  | (-1.676 to 3.361) | (-1.330 to 2.736) | (-5.617 to 10.09) | (-4.508 to 7.574) |
| With hypertension | 2.470 | 1.996 | 7.216 | 5.220 |
|  | (-4.927 to 9.866) | (-3.764 to 7.757) | (-18.31 to 32.75) | (-15.26 to 25.70) |
| Difference<sup>b</sup> | 1.627 | 1.293 | 4.980 | 3.687 |
|  | (-3.268 to 6.522) | (-2.449 to 5.034) | (-12.93 to 22.89) | (-11.04 to 18.41) |
| Observations | 1 331 963 | 1 211 127 | 120 836 | 1 331 963 |
| **Productivity associated costs<sup>c</sup>** |  |  |  |  |
| **Sick absence costs, \$** | | | | |
| Without hypertension | 534.0 | 487.3 | 947.5 | 460.2*** |
|  | (521.0 to 547.0) | (475.4 to 499.2) | (885.2 to 1010) | (397.9 to 522.5) |
| With hypertension | 831.3 | 788.4 | 1212 | 423.8*** |
|  | (790.1 to 872.5) | (746.8 to 829.9) | (1056 to 1368) | (264.3 to 583.3) |
| Difference<sup>b</sup> | **297.3\*\*\*** | **301.0\*\*\*** | **264.6\*\*** | -36.43 |
|  | **(254.1 to 340.6)** | **(257.9 to 344.2)** | **(96.50 to 432.7)** | **(-207.7 to 134.9)** |
| Observations | 139 490 | 125 345 | 14 145 | 139 490 |
| **STD costs, \$** | | | | |
| Without hypertension | 469.0 | 402.6 | 1149 | 746.0*** |
|  | (454.6 to 483.3) | (391.2 to 414.1) | (1084 to 1213) | (686.7 to 805.3) |
| With hypertension | 1100 | 996.9 | 2156 | 1160*** |
|  | (1056 to 1144) | (956.0 to 1038) | (1946 to 2367) | (951.8 to 1367) |
| Difference<sup>b</sup> | **631.0\*\*\*** | **594.2\*\*\*** | **1008\*\*\*** | **413.6\*\*\*** |
|  | **(591.4 to 670.7)** | **(556.9 to 631.6)** | **(800.3 to 1215)** | **(205.0 to 622.2)** |
| Observations | 1 355 064 | 1 234 579 | 120 485 | 1 355 064 |
| **LTD costs, \$** | | | | |
| Without hypertension | 123.8 | 103.4 | 328.8 | 225.4 |
|  | (-190.4 to 438.1) | (-150.3 to 357.1) | (-650.8 to 1308) | (-527.9 to 978.7) |
| With hypertension | 363.1 | 293.5 | 1061 | 767.7 |
|  | (-559.8 to 1286) | (-425.3 to 1012) | (-2124 to 4246) | (-1787 to 3322) |
| Difference<sup>b</sup> | 239.3 | 190.1 | 732.4 | 542.3 |
|  | (-371.5 to 850.1) | (-276.8 to 657.0) | (-1,502 to 2,967) | (-1,294 to 2,379) |
| Observations | 1 331 963 | 1 211 127 | 120 836 | 1 331 963 |

Abbreviations: LTD, long-term disability; STD, short-term disability.
\*\*\* p<0.001, \*\* p<0.01, \* p<0.05
<sup>a</sup> A negative binomial model was used for the number of sick absences, STD, and LTD. A generalized linear model for costs associated with sick absences, STD, and LTD. All estimates were average marginal effects with 95% confidence interval (CI), adjusting patients' age groups, sex, urbanicity of residence, Census regions, and comorbidities.
<sup>b</sup> The differences in the predicted values, ie, average marginal effects associated with hypertension.
<sup>c</sup> The productivity costs associated with sick absences, STD, and LTD were obtained by multiplying the number of hours of sick absences, STD, and LTD by the average hourly wage in 2021. Adjustments for absences associated with short-term (70% of hourly wage) and long-term (60% of hourly wage) disability were made. The average hourly wage for all employees on private nonfarm payrolls was derived from the US Bureau of Labor Statistics.

The sensitivity analyses using a zero-inflated negative binomial and a two-part model are presented in eTable 2. The magnitudes and directions were similar to results from the main analyses, except the number and costs of LTD by COVID-19 diagnosis status became statistically significant.

eTables 3 and 4 presented the same results as Tables 2 and 3 using 2020 data. The total medical costs associated with hypertension were $8961 (95% CI, $8478-$9445; *P* < 0.001) in 2020 (eTable 3), very similar to the 2021 estimate of $8723 using 2021 data (Table 2). On the other hand, the differences in total excess medical costs associated with hypertension by COVID-19 diagnosis status were much higher in 2020 estimates ($18 167; 95% CI, $14 320-$22 014; *P* < 0.001) than 2021 estimates ($6117; 95% CI, 4780-7453) (Table 2). The differences in excess inpatient admission associated with hypertension by COVID-19 diagnosis status were 0.986 per patient (95% CI, 0.825-1.147; *P* < 0.001) (eTable 3), more than 3 times higher than the 0.293 estimate using 2021 data (Table 2). Similarly, the differences in the number of excess ED visits and outpatient prescription drugs were higher in 2020 than in 2021. The estimated differences of excess productivity losses, and costs associated with hypertension by COVID-19 diagnosis in 2020 (eTable 4), were similar to estimates in 2021.

## DISCUSSION

This is the first study to assess the excess medical costs, health care utilization, and productivity-related outcomes and costs associated with hypertension by COVID-19 diagnosis. In 2021, adults aged 18-64 years with employer-sponsored private insurance experienced an estimated excess medical cost of $8723 per patient associated with hypertension. Meanwhile, the combined productivity loss attributable to hypertension resulting from sick absences and STD was $928 per patient. Furthermore, patients with hypertension who had a COVID-19 diagnosis exhibited significantly higher medical costs compared to those without COVID-19. Overall, the excess medical cost associated with hypertension was $6117 higher for individuals with a COVID-19 diagnosis in comparison to those without a COVID-19 diagnosis. Furthermore, a COVID-19 diagnosis was associated with a $414 increase in hypertension-associated excess STD-related costs, whereas sick absences and LTD-related productivity losses were not significantly different for persons with and without COVID-19. The higher medical cost associated with hypertension among patients with a COVID-19 diagnosis was primarily driven by increased ED visits, inpatient admissions, and outpatient prescription drugs.

Given the interdependent relationship between hypertension and COVID-19 in our regression models, our findings can also be interpreted as excess costs, utilization and productivity losses associated with COVID-19 diagnosis among individuals with and without hypertension. For instance, total medical costs associated with COVID-19 can be interpreted as being significantly higher among persons with hypertension compared to those without.

Similarly, other health care utilization and productivity outcomes indicated considerably higher associations with COVID-19 diagnosis among individuals with hypertension compared to those without.

Our findings are consistent with prior research, indicating elevated number of ED visits, inpatient and outpatient treatment, and medication use among adults with hypertension compared to those without.^5,6^ However, our estimated medical cost associated with hypertension ($8723 per patient per year) significantly surpasses prepandemic estimates derived from national household surveys, which were typically $1500-$2500 per person annually.^5–7^ Several factors may underlie these discrepancies. First, the focus of our study on the COVID-19 period may inherently differ from prepandemic studies, in terms of health care utilization and medical costs. Second, our study may differ from prior research that encompassed individuals across various insurance types. This difference may explain variations in costs between privately insured employed adults in our study, and previous studies’ populations, which likely also included those covered by public insurance as well as uninsured population. Third, previous studies using survey data identified individuals with lifetime hypertension, whereas our analysis, based on claims databases, focused on individuals diagnosed with hypertension within the study year.

Adults with documented lifetime hypertension in survey databases may have received ongoing management and control for their condition, potentially resulting in lower recorded hypertension-related medical costs compared to those diagnosed within our study sample. And fourth, our stringent criteria for identifying individuals with hypertension—requiring them to have at least 1 inpatient admission or 2 outpatient visits with a hypertension diagnosis—might have inadvertently excluded individuals with less severe hypertension.

Preexisting hypertension may increase the risk of severe COVID-19 symptoms; likewise, COVID-19 may be associated with subsequent onset or exacerbation of hypertension.^20–22,39^ For example, existing evidence has linked hypertension with increased hyperinflammatory responses following a SARS-CoV-19 infection, which may contribute to a more severe trajectory of COVID-19.^20^ In a study involving more than 5500 inpatients with COVID-19 who had no history of hypertension, 21% developed new-onset hypertension during hospitalization.^22^ The potential correlation between hypertension and more severe COVID-19 symptoms may in part explain our findings that hypertension-related health care utilization and costs are higher among persons with a COVID-19 diagnosis.^24,40^

Also, in our sensitivity analysis, we observed significantly higher medical costs associated with hypertension among those diagnosed with COVID-19 in 2020 compared to 2021. The introduction of mRNA-based COVID-19 vaccines, proven safe and effective for individuals with hypertension,^41^ might have played a role in the reduction of hypertension-related medical costs in 2021 vs 2020.^42^ Further exploration may be required to understand the temporal impact of vaccination on hypertension disease severity and medical costs. Understanding the evolving dynamics between COVID-19 disease management, vaccination programs, and associated medical costs—especially among those with poor baseline health and/or comorbidities like hypertension—may help assess the broader implications of COVID-19 interventions on health care costs and outcomes.

Lastly, prior research has consistently demonstrated the substantial impact of hypertension on productivity, contributing to increased work absences, compromised on-the-job productivity, and heightened disabilities.^10^ Our study further contributes to this literature by unveiling the compounded effect of a COVID-19 diagnosis on productivity losses associated with hypertension. This finding illuminates the intricate interplay between COVID-19 and hypertension, encouraging a reevaluation of the broader economic repercussions of the pandemic. In this context, it becomes crucial for future investigations to not only account for the direct medical and productivity costs linked to COVID-19 treatment but also to consider the additional economic burden in managing concurrent chronic conditions such as hypertension.

Understanding and addressing these intertwined health care challenges are pivotal for devising comprehensive strategies aimed at mitigating the multidimensional impacts of global health crises, exemplified by the ongoing challenges posed by COVID-19.

### Limitation

This study is subject to several limitations. First, the MarketScan Commercial Database, sourced from large employers, represents a convenience sample, limiting the generalizability of findings to privately insured individuals with payroll information. Consequently, the study findings cannot be generalized to the broader spectrum of privately insured US adults. Second, the findings might not accurately represent individuals lacking health insurance or covered by public insurance programs like Medicare and Medicaid. Notably, one of our primary outcomes— productivity losses—is contingent upon data from employers’ payroll systems, thereby excluding persons without employment or those covered by public insurance. Third, our study focused solely on surviving individuals with hypertension and COVID-19, precluding the assessment of costs and productivity due to mortality. Fourth, we were unable to ascertain the severity levels of hypertension and COVID-19, which could have distinct effects on the outcomes, potentially leading to an underestimation of our results. Lastly, while emerging evidence links hypertension with an elevated risk of Long COVID symptoms,^43^ our cross-sectional data precluded an evaluation of the longitudinal impact of a COVID-19 diagnosis on hypertension-related health care utilization and economic costs.

## CONCLUSIONS

The economic burden of hypertension, in terms of medical costs and productivity loss, is substantial. Our study reveals that individuals diagnosed with COVID-19 incurred even higher excess medical costs, health care utilization, and productivity losses attributable to hypertension. These findings emphasize the importance of factoring in costs of chronic comorbidities in assessing the overall economic burden of COVID-19. These estimates can help assess the economic impact (eg, costs averted) of interventions effective in preventing COVID-19 among individuals with preexisting hypertension.

## Data Availability

The authors cannot share the data publicly due to the data user agreement, but the program codes are available upon request to the corresponding author.

## Acknowledgements

We thank Michael Schooley, Janet Wright, and Fátima Coronado (Centers for Disease Control and Prevention) for their guidance, suggestions, and manuscript review. We would like to thank Moira Urich (the Centers for Disease Control and Prevention) for review of this manuscript.

---

**eTable 1.**
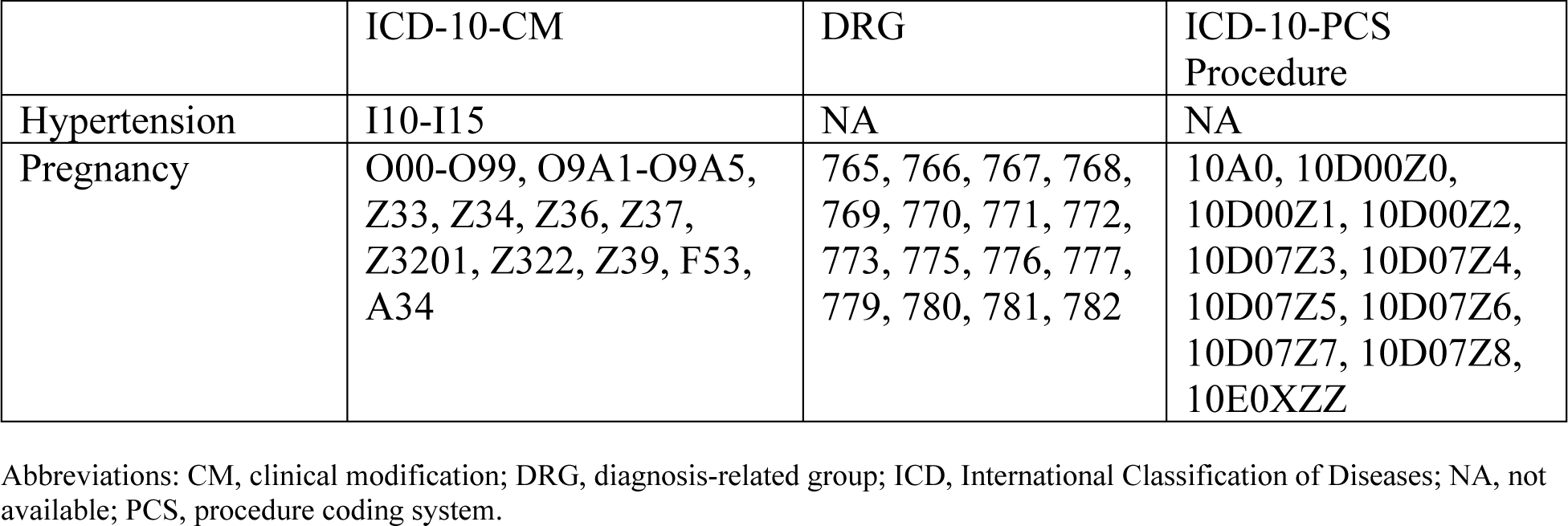
The ICD-10-CM for Hypertension and ICD-10-CM, DRG, and ICD-10-PCS Procedure Codes Used for Pregnancy Exclusion.

**eTable 2.** Productivity Losses and Costs Associated With Hypertension Using a Zero-Inflated Negative Binomial and a Two-Part Model, 2021^a^.

|  | All | Without COVID-19 diagnosis | With COVID-19 diagnosis | With COVID-19 vs without COVID-19 |
| --- | --- | --- | --- | --- |
| **Sick absences (days)** |  |  |  |  |
| Without hypertension | 2.163 | 2.028 | 3.360 | 1.332*** |
|  | (2.114 to 2.212) | (1.979 to 2.076) | (3.200 to 3.520) | (1.173 to 1.492) |
| With hypertension | 3.180 | 3.069 | 4.163 | 1.094*** |
|  | (3.018 to 3.342) | (2.898 to 3.240) | (3.775 to 4.550) | (0.685 to 1.503) |
| Difference<sup>b</sup> | 1.017*** | 1.041*** | 0.803*** | -0.239 |
|  | (0.845 to 1.190) | (0.860 to 1.223) | (0.387 to 1.219) | (-0.676 to 0.199) |
| Observations | 139 490 | 125 345 | 14 145 | 139 490 |
| **STD (days)** |  |  |  |  |
| Without hypertension | 2.958 | 3.015 | 2.381 | -0.633*** |
|  | (2.924 to 2.993) | (2.978 to 3.051) | (2.331 to 2.432) | (-0.691 to -0.576) |
| With hypertension | 3.398 | 3.424 | 3.130 | -0.293*** |
|  | (3.337 to 3.459) | (3.359 to 3.488) | (3.032 to 3.229) | (-0.400 to -0.187) |
| Difference<sup>b</sup> | **0.439\*\*\*** | **0.409\*\*\*** | **0.749\*\*\*** | **0.340\*\*\*** |
|  | **(0.375 to 0.504)** | **(0.340 to 0.478)** | **(0.641 to 0.857)** | **(0.219 to 0.461)** |
| Observations | 1 355 064 | 1 234 579 | 120 485 | 1 355 064 |
| **LTD (days)** |  |  |  |  |
| Without hypertension | 0.456 | 0.459 | 0.427 | -0.0318 |
|  | (0.426 to 0.486) | (0.427 to 0.491) | (0.384 to 0.470) | (-0.0803 to 0.0167) |
| With hypertension | 0.469 | 0.464 | 0.514 | 0.0504 |
|  | (0.440 to 0.498) | (0.435 to 0.493) | (0.448 to 0.581) | (-0.0155 to 0.116) |
| Difference<sup>b</sup> | 0.0125 | 0.00508 | 0.0873* | **0.0822\*** |
|  | (-0.0206 to 0.0456) | (-0.0303 to 0.0404) | (0.0122 to 0.162) | **(0.000322 to 0.164)** |
| Observations | 1 331 963 | 1 211 127 | 120 836 | 1 331 963 |
| **Productivity associated costs<sup>c</sup>** |  |  |  |  |
| **Sick absence costs, \$** | | | | |
| Without hypertension | 533.9 | 488.1 | 940.0 | 451.9*** |
|  | (522.0 to 545.9) | (476.6 to 499.6) | (887.8 to 992.2) | (399.4 to 504.5) |
| With hypertension | 826.4 | 783.6 | 1205 | 421.4*** |
|  | (788.9 to 863.8) | (745.2 to 822.0) | (1077 to 1333) | (290.0 to 552.9) |
| Difference<sup>b</sup> | **292.4\*\*\*** | **295.5\*\*\*** | **265.0\*\*\*** | -30.50 |
|  | **(252.5 to 332.3)** | **(255.0 to 336.0)** | **(126.4 to 403.7)** | (-172.2 to 111.2) |
| Observations | 139 490 | 125 345 | 14 145 | 139 490 |
| **STD costs, \$** | | | | |
| Without hypertension | 444.6 | 390.6 | 997.6 | 607.0*** |
|  | (439.2 to 450.0) | (385.3 to 396.0) | (974.4 to 1021) | (583.4 to 630.6) |
| With hypertension | 1014 | 931.4 | 1857 | 926.0*** |
|  | (992.5 to 1035) | (909.7 to 953.1) | (1785 to 1930) | (852.3 to 999.8) |
| Difference<sup>b</sup> | **569.2\*\*\*** | **540.8\*\*\*** | **859.9\*\*\*** | **319.1\*\*\*** |
|  | **(546.7 to 591.7)** | **(518.0 to 563.6)** | **(783.3 to 936.4)** | **(241.5 to 396.6)** |
| Observations | 1 355 064 | 1 234 579 | 120 485 | 1 355 064 |
| **LTD costs, \$** | | | | |
| Without hypertension | 52.72 | 48.97 | 90.28 | 41.30*** |
|  | (49.19 to 56.24) | (45.47 to 52.47) | (75.28 to 105.3) | (26.09 to 56.52) |
| With hypertension | 133.2 | 123.2 | 232.8 | 109.6*** |
|  | (121.4 to 144.9) | (111.4 to 135.1) | (189.7 to 275.9) | (65.82 to 153.3) |
| Difference<sup>b</sup> | **80.47\*\*\*** | **74.28\*\*\*** | **142.5\*\*\*** | **68.25\*\*** |
|  | **(67.74 to 93.19)** | **(61.58 to 86.98)** | **(96.45 to 188.6)** | **(21.88 to 114.6)** |
| Observations | 1 331 963 | 1 211 127 | 120 836 | 1 331 963 |

Abbreviations: LTD, long-term disability; STD, short-term disability.
\*\*\* $P < 0.001$ , \*\* $P < 0.01$ , \* $P < 0.05$
<sup>a</sup> This table provides the same dependent variables with Table 3 but with different models for robustness check. A zero-inflated negative binomial model was used for number of sick absences, STD, and LTD. A two-part model was used, where the first part is a logistic regression model, and the second part is a generalized linear model for costs associated with sick absences, STD, and LTD. All estimates were average marginal effects with 95% confidence interval (CI), adjusting patients' age groups, sex, urbanicity of residence, Census regions, and comorbidities.
<sup>b</sup> The differences in the predicted values, ie, average marginal effects associated with hypertension.
<sup>c</sup> The productivity costs associated with sick absences, STD, and LTD were obtained by multiplying the number of hours of sick absences, STD, and LTD by the average hourly wage in 2021. Adjustments for absences associated with short-term (70% of hourly wage) and long-term (60% of hourly wage) disability were made. The average hourly wage for all employees on private nonfarm payrolls was derived from the US Bureau of Labor Statistics.

**eTable 3.**
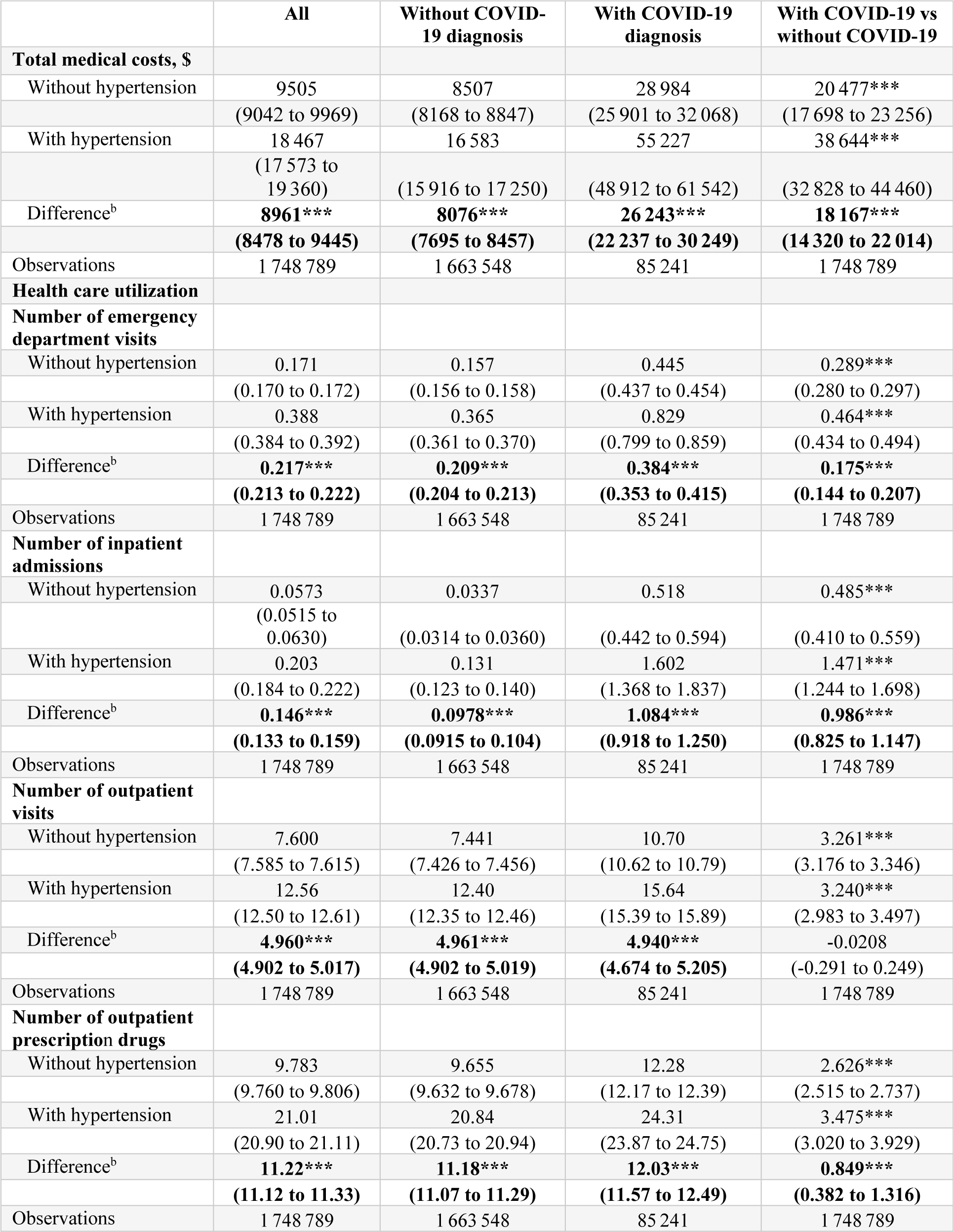
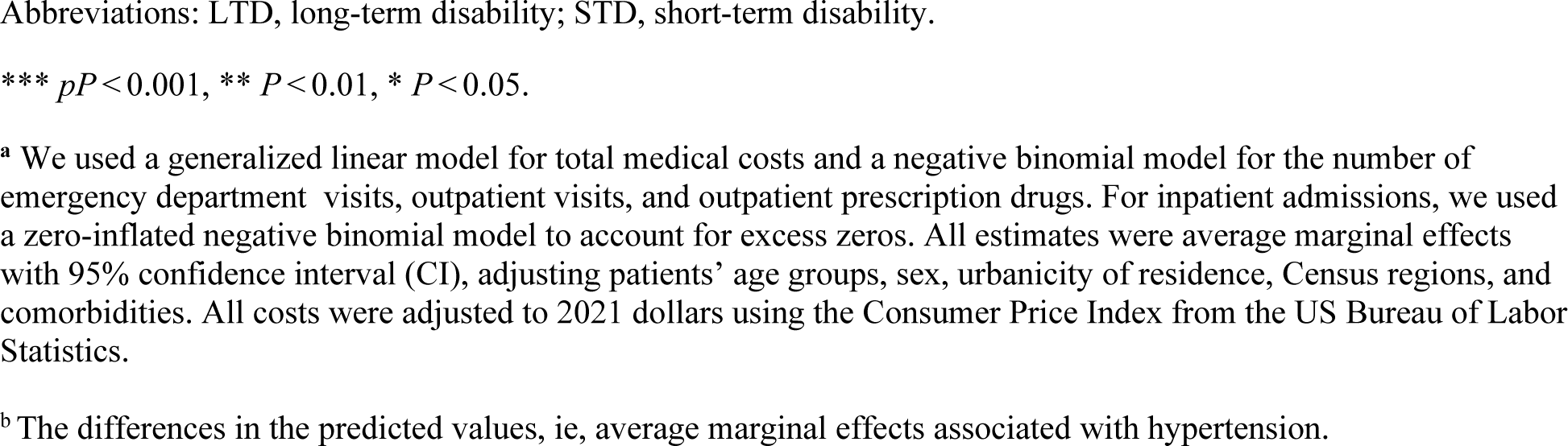
Supplementary Analysis Using 2020 Data—Total Medical Costs and Health Care Utilization Associated With Hypertension^a^.

|  | All | Without COVID-19 diagnosis | With COVID-19 diagnosis | With COVID-19 vs without COVID-19 |
| --- | --- | --- | --- | --- |
| **Total medical costs, \$** | | | | |
| Without hypertension | 9505 | 8507 | 28 984 | 20 477*** |
|  | (9042 to 9969) | (8168 to 8847) | (25 901 to 32 068) | (17 698 to 23 256) |
| With hypertension | 18 467 | 16 583 | 55 227 | 38 644*** |
|  | (17 573 to 19 360) | (15 916 to 17 250) | (48 912 to 61 542) | (32 828 to 44 460) |
| Difference<sup>b</sup> | **8961\*\*\*** | **8076\*\*\*** | **26 243\*\*\*** | **18 167\*\*\*** |
|  | **(8478 to 9445)** | **(7695 to 8457)** | **(22 237 to 30 249)** | **(14 320 to 22 014)** |
| Observations | 1 748 789 | 1 663 548 | 85 241 | 1 748 789 |
| **Health care utilization** |  |  |  |  |
| **Number of emergency department visits** |  |  |  |  |
| Without hypertension | 0.171 | 0.157 | 0.445 | 0.289*** |
|  | (0.170 to 0.172) | (0.156 to 0.158) | (0.437 to 0.454) | (0.280 to 0.297) |
| With hypertension | 0.388 | 0.365 | 0.829 | 0.464*** |
|  | (0.384 to 0.392) | (0.361 to 0.370) | (0.799 to 0.859) | (0.434 to 0.494) |
| Difference<sup>b</sup> | **0.217\*\*\*** | **0.209\*\*\*** | **0.384\*\*\*** | **0.175\*\*\*** |
|  | **(0.213 to 0.222)** | **(0.204 to 0.213)** | **(0.353 to 0.415)** | **(0.144 to 0.207)** |
| Observations | 1 748 789 | 1 663 548 | 85 241 | 1 748 789 |
| **Number of inpatient admissions** |  |  |  |  |
| Without hypertension | 0.0573 | 0.0337 | 0.518 | 0.485*** |
|  | (0.0515 to 0.0630) | (0.0314 to 0.0360) | (0.442 to 0.594) | (0.410 to 0.559) |
| With hypertension | 0.203 | 0.131 | 1.602 | 1.471*** |
|  | (0.184 to 0.222) | (0.123 to 0.140) | (1.368 to 1.837) | (1.244 to 1.698) |
| Difference<sup>b</sup> | **0.146\*\*\*** | **0.0978\*\*\*** | **1.084\*\*\*** | **0.986\*\*\*** |
|  | **(0.133 to 0.159)** | **(0.0915 to 0.104)** | **(0.918 to 1.250)** | **(0.825 to 1.147)** |
| Observations | 1 748 789 | 1 663 548 | 85 241 | 1 748 789 |
| **Number of outpatient visits** |  |  |  |  |
| Without hypertension | 7.600 | 7.441 | 10.70 | 3.261*** |
|  | (7.585 to 7.615) | (7.426 to 7.456) | (10.62 to 10.79) | (3.176 to 3.346) |
| With hypertension | 12.56 | 12.40 | 15.64 | 3.240*** |
|  | (12.50 to 12.61) | (12.35 to 12.46) | (15.39 to 15.89) | (2.983 to 3.497) |
| Difference<sup>b</sup> | **4.960\*\*\*** | **4.961\*\*\*** | **4.940\*\*\*** | -0.0208 |
|  | **(4.902 to 5.017)** | **(4.902 to 5.019)** | **(4.674 to 5.205)** | **(-0.291 to 0.249)** |
| Observations | 1 748 789 | 1 663 548 | 85 241 | 1 748 789 |
| **Number of outpatient prescription drugs** |  |  |  |  |
| Without hypertension | 9.783 | 9.655 | 12.28 | 2.626*** |
|  | (9.760 to 9.806) | (9.632 to 9.678) | (12.17 to 12.39) | (2.515 to 2.737) |
| With hypertension | 21.01 | 20.84 | 24.31 | 3.475*** |
|  | (20.90 to 21.11) | (20.73 to 20.94) | (23.87 to 24.75) | (3.020 to 3.929) |
| Difference<sup>b</sup> | **11.22\*\*\*** | **11.18\*\*\*** | **12.03\*\*\*** | **0.849\*\*\*** |
|  | **(11.12 to 11.33)** | **(11.07 to 11.29)** | **(11.57 to 12.49)** | **(0.382 to 1.316)** |
| Observations | 1 748 789 | 1 663 548 | 85 241 | 1 748 789 |

Abbreviations: LTD, long-term disability; STD, short-term disability.
\*\*\* $pP < 0.001$ , \*\* $P < 0.01$ , \* $P < 0.05$ .
<sup>a</sup> We used a generalized linear model for total medical costs and a negative binomial model for the number of emergency department visits, outpatient visits, and outpatient prescription drugs. For inpatient admissions, we used a zero-inflated negative binomial model to account for excess zeros. All estimates were average marginal effects with 95% confidence interval (CI), adjusting patients' age groups, sex, urbanicity of residence, Census regions, and comorbidities. All costs were adjusted to 2021 dollars using the Consumer Price Index from the US Bureau of Labor Statistics.
<sup>b</sup> The differences in the predicted values, ie, average marginal effects associated with hypertension.

**eTable 4.** Supplementary Analysis Using 2020 Data—Productivity Losses and Costs Associated With Hypertension^a^.

|  | All | Without COVID-19 diagnosis | With COVID-19 diagnosis | With COVID-19 vs without COVID-19 |
| --- | --- | --- | --- | --- |
| **Sick absences (days)** |  |  |  |  |
| Without hypertension | 1.843 | 1.790 | 2.780 | 0.990*** |
|  | (1.817 to 1.869) | (1.765 to 1.815) | (2.626 to 2.934) | (0.836 to 1.144) |
| With hypertension | 2.704 | 2.639 | 3.841 | 1.202*** |
|  | (2.621 to 2.786) | (2.555 to 2.723) | (3.437 to 4.246) | (0.793 to 1.612) |
| Difference<sup>b</sup> | **0.860\*\*\*** | **0.849\*\*\*** | **1.061\*\*\*** | 0.212 |
|  | **(0.772 to 0.948)** | **(0.761 to 0.937)** | **(0.629 to 1.494)** | (-0.225 to 0.650) |
| Observations | 222 101 | 210 181 | 11 920 | 222 101 |
| **STD (days)** |  |  |  |  |
| Without hypertension | 2.583 | 2.367 | 6.805 | 4.438*** |
|  | (2.485 to 2.682) | (2.290 to 2.444) | (6.073 to 7.537) | (3.750 to 5.126) |
| With hypertension | 6.155 | 5.801 | 13.06 | 7.261*** |
|  | (5.846 to 6.464) | (5.518 to 6.084) | (10.81 to 15.31) | (5.048 to 9.475) |
| Difference<sup>b</sup> | **3.572\*\*\*** | **3.434\*\*\*** | **6.258\*\*\*** | **2.824\*** |
|  | **(3.292 to 3.851)** | **(3.169 to 3.699)** | **(4.108 to 8.408)** | **(0.674 to 4.973)** |
| Observations | 1 521 915 | 1 447 756 | 74 159 | 1 521 915 |
| **LTD (days)** |  |  |  |  |
| Without hypertension | 0.439 | 0.441 | 0.397 | -0.0447 |
|  | (0.412 to 0.466) | (0.413 to 0.469) | (0.335 to 0.459) | (-0.109 to 0.0198) |
| With hypertension | 0.447 | 0.451 | 0.378 | -0.0734* |
|  | (0.405 to 0.490) | (0.407 to 0.495) | (0.316 to 0.440) | (-0.138 to -0.00932) |
| Difference<sup>b</sup> | 0.00832 | 0.00976 | -0.0190 | -0.0287 |
|  | (-0.0322 to 0.0488) | (-0.0323 to 0.0518) | (-0.103 to 0.0651) | (-0.119 to 0.0620) |
| Observations | 1 470 046 | 1 396 791 | 73 255 | 1 470 046 |
| **Productivity associated costs<sup>c</sup>** |  |  |  |  |
| **Sick absence costs, \$** | | | | |
| Without hypertension | 453.2 | 440.1 | 684.8 | 244.7*** |
|  | (445.3 to 461.1) | (432.4 to 447.8) | (637.1 to 732.4) | (197.0 to 292.4) |
| With hypertension | 665.1 | 649.2 | 946.1 | 296.9*** |
|  | (639.5 to 690.7) | (623.4 to 675.0) | (820.0 to 1,072) | (169.3 to 424.6) |
| Difference<sup>b</sup> | **211.9\*\*\*** | **209.1\*\*\*** | **261.3\*\*\*** | 52.25 |
|  | **(184.7 to 239.0)** | **(181.8 to 236.3)** | **(126.4 to 396.2)** | (-83.98 to 188.5) |
| Observations | 222 101 | 210 181 | 11 920 | 222 101 |
| **STD costs, \$** | | | | |
| Without hypertension | 444.7 | 407.5 | 1172 | 764.2*** |
|  | (432.2 to 457.3) | (397.8 to 417.2) | (1079 to 1265) | (677.0 to 851.4) |
| With hypertension | 1060 | 998.7 | 2249 | 1251*** |
|  | (1020 to 1099) | (962.9 to 1035) | (1964 to 2535) | (969.9 to 1531) |
| Difference<sup>b</sup> | **614.9\*\*\*** | **591.2\*\*\*** | **1078\*\*\*** | **486.3\*\*\*** |
|  | **(579.6 to 650.3)** | **(557.7 to 624.8)** | **(805.0 to 1350)** | **(213.9 to 758.7)** |
| Observations | 1 521 915 | 1 447 756 | 74 159 | 1 521 915 |
| **LTD costs, \$** | | | | |
| Without hypertension | 97.21 | 90.79 | 219.7 | 128.9 |
|  | (-36.18 to 230.6) | (-29.58 to 211.2) | (-213.0 to 652.4) | (-205.8 to 463.6) |
| With hypertension | 281.2 | 263.3 | 622.6 | 359.4 |
|  | (-102.9 to 665.2) | (-84.64 to 611.1) | (-677.6 to 1923) | (-686.2 to 1405) |
| Difference<sup>b</sup> | 184.0 | 172.5 | 403.0 | 230.5 |
|  | (-70.17 to 438.1) | (-58.48 to 403.4) | (-528.1 to 1334) | (-556.1 to 1017) |
| Observationss | 1 470 046 | 1 396 791 | 73 255 | 1 470 046 |

Abbreviations: LTD, long-term disability; STD, short-term disability.
\*\*\* $Pp < 0.001$ , \*\* $P < 0.01$ , \* $P < 0.05$ .
<sup>a</sup> A negative binomial model was used for the number of sick absences and STD. For number of LTD, we used a zero-inflated negative binomial model. A generalized linear model for costs associated with sick absences, STD, and LTD. All estimates were average marginal effects with 95% confidence interval (CI), adjusting patients' age groups, sex, urbanicity of residence, Census regions, and comorbidities. All costs were adjusted to 2021 dollars using the Consumer Price Index from the US Bureau of Labor Statistics.
<sup>b</sup> The differences in the predicted values, ie, average marginal effects associated with hypertension.
<sup>c</sup> The productivity costs associated with sick absences, STD, and LTD were obtained by multiplying the number of hours of sick absences, STD, and LTD by the average hourly wage in 2020. Adjustments for absences associated with short-term (70% of hourly wage) and long-term (60% of hourly wage) disability were made. The average hourly wage for all employees on private nonfarm payrolls was derived from the US Bureau of Labor Statistics.

